# Machine learning and the labour market: A portrait of occupational and worker inequities in Canada

**DOI:** 10.1101/2024.06.12.24308855

**Authors:** Arif Jetha, Qing Liao, Faraz Vahid Shahidi, Viet Vu, Aviroop Biswas, Brendan Smith, Peter Smith

## Abstract

**Introduction:** Machine learning (ML) is increasingly used by Canadian workplaces. Concerningly, the impact of ML may be inequitable and disrupt social determinants of health. The aim of this study is to estimate the number of workers in occupations highly exposed to ML and describe differences in ML exposure represents according to occupational and worker sociodemographic factors.

**Methods:** Canadian occupations were scored according to the extent to which they were made up of job tasks that could be performed by ML. Eight years of data from Canada’s Labour Force Survey were pooled and the number of Canadians in occupations with high or low exposed to machine learning were estimated. The relationship between gender, hourly wages, educational attainment and occupational job skills, experience and training requirements and ML exposure was examined using stratified logistic regression models.

**Results:** Approximately, 1.9 million Canadians are working in occupations with high ML exposure and 744,250 workers were employed in occupations with low ML exposure. Women were more likely to be employed in occupations with high ML exposure than men. Workers with greater educational attainment and in occupations with higher wages and greater job skills requirements were more likely to experience high ML exposure. Women, especially those with less educational attainment and in jobs with greater job skills, training and experience requirements, were disproportionately exposed to ML.

**Conclusion:** ML has the potential to widen inequities in the working population. Disadvantaged segments of the workforce may be most likely to be employed in occupations with high ML exposure. ML may have a gendered effect and disproportionately impact certain groups of women when compared to men. We provide a critical evidence base to develop strategic responses that ensure inclusion in a working world where ML is commonplace.

**KEY MESSAGES:** *What is already known on this topic:*

- The Canadian labour market is undergoing an artificial intelligence (AI) revolution that has the potential to have widespread impact on a range of occupations and worker groups.
- It is unclear how which the adoption of machine learning (ML), an AI subfield, within the working world might contribute to inequities within the labour market.

*What this study adds:*

- Segments of the workforce which have been previously disadvantaged may be most likely to work in occupations most likely to be affected by ML.
- ML may have a gendered effect and disproportionately impact some groups of women when compared to men.

*How this study might affect research, practice or policy:*

Findings can inform targeted policies and programs that optimize the economic benefits of ML while addressing disparities that can emerge because of the adoption of the technology on workers.

## INTRODUCTION

Industrialized labour market contexts are increasingly shaped by inequity which are known to influence disparities in the health and quality of life of workers^1^. The rapid adoption of artificial intelligence (AI) has the potential to drive innovation and raise productivity. AI can also contribute to the polarization of the labour market. To date, most studies on the impact of AI focus on aggregate labour market outcomes. There is a lack of descriptive research which examines to examine how AI may differentially impact workers according to occupational or sociodemographic characteristics. Such research allows researchers and policymakers to identify groups of workers who may be most or least likely to be affected by the technology, and who may benefit from direct policy and programmatic support. We use a novel analytical approach and national labour force data to estimate the number of Canadian workers in occupations with high or low machine learning exposure. We then focus on describing the occupational and worker sociodemographic characteristics associated with high or low occupational exposure to ML.

Employment is a critical social determinant of health that provides income and resources that offer pathways to better health ^1^. In Canada, like many other industrialized contexts, the labour market can be segregated according to occupational and sociodemographic factors ^2^ ^3^. Technological transformations have not only been a significant catalyst for economic change within the labour market but have also created both opportunities and barriers for different groups of workers. Previous of periods of technological transformation have increased the likelihood of labour substitution, contributing to partial or full job displacement for workers in certain occupations and potentially eroding working conditions for those who remain employed^4–6^. Workers better positioned in the labour market, including those with more educational attainment and in better paying and higher-skilled jobs have been more likely to capitalize on economic advantage stemming from past technological transformations (e.g., wage growth and access to jobs with better quality). In comparison, workers from socially and economically disadvantaged groups have been adversely affected from past technological transformations and including experiencing job displacement or an erosion to job quality^6^ ^7^.

Research on the automation of work provides an example of how a technological transformation can widen inequities. Past periods of automation have shown that job tasks most likely to be performed by computer software, robots or other digital technologies included those that were routine, structured or repetitive that are most common in administrative, manufacturing or service-oriented occupations ^5^ ^6^ ^8^. Research also indicates that worker sociodemographic and occupational characteristics were associated with the automation of work; workers with greater educational attainment and job skills, experience and training requirements and those that earn greater wages may be more likely to work in jobs with variable or cognitively demanding job tasks that were less susceptible to automation^9–11^. Studies of past waves of automation have also indicated gender difference^12^. Industries to which men are employed (e.g., repair, construction, and transportation) may be more likely to characterized by repetitive job tasks that are most suitable to being performed by robots or machines^13^ and the requirement for social and emotional job skills in occupations disproportionately held by women (e.g., education, healthcare, and administration) could be protective against automation ^14^. Other research suggests that women may be less likely to be employed in occupations with managerial or professional occupations or are underrepresented in science, technology, engineering and mathematics jobs that may benefit increased productivity related to the introduction of new digital technologies ^12^ ^15^ ^16^.

Our study focuses on the implications of machine learning (ML), an AI subfield, for the employment and equity of the Canadian working population. ML involves the application of statistical algorithms to large amounts of data with the aim of making probabilistic inferences about a range of outcomes. ML is being adopted by workplaces to detect patterns in structured or unstructured data, make predictions, generate decisions, or optimize processes ^17^ ^18^. Examples of the use of ML exist across all industries, such as healthcare settings (e.g., use of ML to detect disease in medical imaging ^19^), financial services (e.g., ML-based fraud detection tools ^20^) and the transportation sector (e.g., ML real-time hazard identification in automated vehicles^21^). ML may have a distinct impact on the labour market in ways that are different from past waves of automation ^5^. The capacity for ML to learn, adapt, and generate outputs with increasing independence means that the technology can perform job tasks that are physical or cognitive in nature and are across a broader range of occupations ^17^. It is important to highlight that rapid advancement in ML technologies and its processing power can double the speed of ML every 4 to 9 months, resulting in an evolving learning capacity of the technology and constantly changing impact on workers and workplaces ^5^. Innovations in deep learning, a subset of ML using neural networks (i.e., algorithms with structures inspired by human brain function) are also expected to enhance ML performance and its capacity to automate a greater range of job tasks including those that involve image and speech recognition and predictive analytics which could have significant implications for the working population.

Studies examining the extent to which job tasks that make up different occupations are suitable to being performed by ML (*from this point forward referred to as occupational ML exposure*) in the US and Canada suggest that almost all occupations include at least some job tasks that can be performed by ML ^22^. Other research indicates that 19 percent of US occupations may be highly exposed to ML ^23^. Of note, mirroring past waves of automation, no single occupation can be fully performed by ML ^5^. As a result, it is expected that ML is unlikely to be used by workplaces to fully substitute human workers for machines but instead enhance the productivity of certain groups of workers ^5^ ^24^. Unlike previous waves of automation, ML doesn’t only target routine, repetitive and simple tasks, but can contribute to the automation of job tasks that can include planning, learning, reasoning, problem-solving or prediction that tend to cluster in occupations with greater job skills, training and experience requirements and among workers with greater levels of educational attainment ^17^. Accordingly, the impact of ML on the labour market is thought to have a unique and significant impact on occupations and different groups of workers. It is unclear whether occupational ML exposure which may be suitable to performing a greater range of job tasks to may exacerbate or minimize inequities in the Canadian working population.

Describing differences in the labour market related to occupational ML exposure represents an important strategy to build an evidence base that can be used to identify and address emerging occupational and worker inequities and examine how the rapid adoption of the technology can have implications for the social determinants of health. Our study describes divisions in Canada’s labour market related to the ongoing adoption of ML and help to identify groups of workers most affected by the technology. Our research has three main study objectives, which include:

1. Estimating the number of workers in occupations characterized by high or low ML exposure in Canada.
2. Examining how ML exposure differs according to workers’ socioeconomic (e.g., educational attainment, gender) and occupational characteristics (e.g., job skill level requirements, hourly wages).
3. Determining whether gender moderates the association between educational attainment, job skill requirements, hourly wages, and ML exposure.

## METHODS

We conducted a pooled cross-sectional analysis of eight years of Statistics Canada’s Labour Force Survey (LFS; 2013-2019, 2022). The LFS is a nationally-representative cross-sectional monthly survey of the household population (≥15 years of age) with an approximate sample of 100,000 Canadians built using probability sampling procedures ^26^. Excluded from the survey’s coverage are persons who are not currently employed, self-employed, living on reserves and other Indigenous settlements, full-time members of the Canadian Armed Forces, the institutionalized population, and households in extremely remote areas with very low population density. Weighted monthly cycles were combined to produce annual estimates. LFS waves 2020-2021 were excluded from the analysis due to the considerable economic impact of the COVID-19 pandemic which may have significantly skewed study results, given the unequal impacts of COVID-19 on labour market participation across industries and labour force subgroups ^27^.

### Suitability for Machine Learning Measure (SML)

To measure occupational exposure to ML, we utilized the suitability for machine learning (SML) measure developed by Brynjolfsson and colleagues in the US. The SML estimates the extent to which actions and outputs for a specific job task can be learned by a machine. The SML measure offers detailed information about task-level exposure to ML that can be aggregated to study the impact of ML on the economy ^4^. SML scoring was developed using the US O*NET database which is an American occupational information database describing nearly 1,000 occupations and occupation-specific information via standardized job-oriented descriptors and worker-oriented descriptors ^22^. O*NET describes 2,069 direct work activities and 18,156 job tasks categorized across 873 standardized occupations by using the task ratings file ^28^ ^29^.

A 21-question SML rubric was established to evaluate the criteria required for a machine to substitute an occupational task. The rubric consisted of 23 statements that were evaluated on a five-point scale (5 = strongly agree; 1=strongly disagree) and used to score each direct work activity by multiple raters. Average SML scores of direct work activities associated with each job task were calculated to generate a task-level SML measure. Occupation-level SML scores were then produced by taking the weighted average, by importance, of the tasks mapped to eight-digit O*NET codes ^4^ ^5^. Occupations with a high SML value represent those where ML has the greatest potential to transform a job. To estimate SML values among Canadian occupations, we cross-walked O*NET codes to 5-digit Canadian National Occupation Classification Codes ^30^, which were recently updated in 2021 (NOC 2021). Ultimately, 873 eight digits O*NET codes were mapped to 512 NOC codes with matching attributes ^30^.

Out of 512 occupations for which we can assign a SML value, all Canadian occupations had at least some exposure to ML and no occupation was estimated to be completely exposed to ML. Based on the SML values, we then categorized occupations as high ML exposure (top 10 percentile of SML scores, SML score ≥ 3.597) and low ML exposure (bottom 10 percentile of SML scores, SML score ≤3.351).

### Outcome variables

#### High or low occupational ML exposure

SML scores were used to create two binary outcome variables. The first outcome variable was high occupational ML exposure where occupations with an SML score in the top ten percentile were compared to the reference group of occupational SML scores outside of the top ten percentile. The second outcome was low occupational ML exposure where occupations with an SML score in the bottom ten percentile were compared to the reference group of occupational SML scores outside of the bottom ten percentile.

### Independent variable

#### Gender

A one-item question asked the respondent, “What is your gender?”. For this study, workers identifying as men or women were included in the analytical sample.

#### Educational attainment

The level of educational attainment of workers was measured using the question, “What is the highest certificate, diploma or degree you have obtained?” Based on responses, a four-level categorical variable was created (some post-secondary or less, trades certification or diploma, college or bachelor’s degree, university degree above a bachelor’s degree).

#### Hourly wages

Hourly wages were measured using the question, “What is your hourly rate of pay?” Based on the participant responses, a four-level categorical variable was created based on hourly earnings using interquartile cutoff points at each survey year. Quartile 1 (Q1) represents earnings ≤25th percentile of hourly earnings; Quartile 2 (Q2) represents earnings > 25th percentile (Q1) and ≤ 50th percentile (Q2); Quartile 3 (Q3) denotes hourly earnings > 50th percentile (Q2) and ≤75th percentile (Q3); and Quartile 4 represents >75th percentile of hourly earnings.

#### Job skill, training and experience requirements

TEER (Training, Education, Experience, and Responsibilities) categorizations were assigned to each Canadian occupation by Statistics Canada, reflecting the nature of education and training requirements, experience required to enter an occupation and the complexity of its responsibilities ^30^. TEER categories included: management occupations (TEER 0), professional occupations (requiring a university degree) (TEER 1), skilled, technical, or supervisory occupations (requiring a college diploma, apprenticeship training or have supervisory roles) (TEER 2), semi-skilled occupations (occupations requiring a college diploma, apprenticeship training or more than six months of on-the-job training) (TEER 3) and general skilled occupations (occupations requiring a high school diploma or several weeks of on-the-job training or need short-term work demonstration and no formal education) (TEER 4/5).

### Covariates

Based on information collected in the LFS several covariates were collected in each survey year including sociodemographic characteristics (age [years], recent immigration status [less than 10 year: yes/no] and province of residence) and work context variables (work hours [hours worked/week], industry [goods producing industries or service producing industries], job permanency [e.g., permanently employed, or non-permanently employed] and unionization [yes/no]).

### Analytical Approach

We produced weighted estimates of the number of workers in Canada in occupations with high ML exposure or low ML exposure. Descriptive statistics (weighted counts [n] and percentages [%]) were utilized to describe worker and workplace characteristics according to high or low ML exposure groups. Univariate logistic regression models were run to examine the associations between independent variables and covariates and the likelihood of employment in a high or low ML exposure occupation (Appendix 1)

Finally, multivariable logistic regressions models were fitted to estimate odds and 95% confidence intervals (95%CI) of employment in high ML exposure occupations and likelihood of employment in low ML exposure occupation. Each set of multivariable models were run using one of three independent variables (educational attainment [model 1], hourly wage [model 2], job skill and training requirements [model 3]). Each set of models were stratified by gender and adjusted for sociodemographic and work context covariates. For each of these models, an unstratified multivariable logistic regression model was constructed, incorporating interaction terms between gender and independent variables as well as covariates. Type III tests were then conducted on these unstratified regression models to determine if there were significant differences in effects between men and women. Pearson’s chi-square adjustment was adopted for overdispersion correction and likelihood ratio tests were used to assess goodness of fit of each model. Analyses were conducted using SAS 9.4 ^31^.

## RESULTS

Overall, 1,902,050 Canadian workers were employed in occupations with high ML exposure where the greatest proportion of job tasks are suitable for ML representing (12 percent of the Canadian workforce). In comparison, about 744,250 workers were employed in occupations with low ML exposure where a smaller proportion of job tasks are suitable for ML (4.7 percent of the Canadian workforce). A full list of occupations categorized as high or low ML exposure is presented on Appendix 2. Women were more likely to be employed in occupations with high ML exposure than men (63.4 percent vs. 36.6 percent). Men were more likely to be employed in occupations with low ML exposure than women (59.9 percent vs. 40.1 percent) (Figure 1). The proportion of Canadian workers in occupations with high or low exposure to ML remained stable over the 8-year measurement period (Appendix 3).

**Figure 1.**
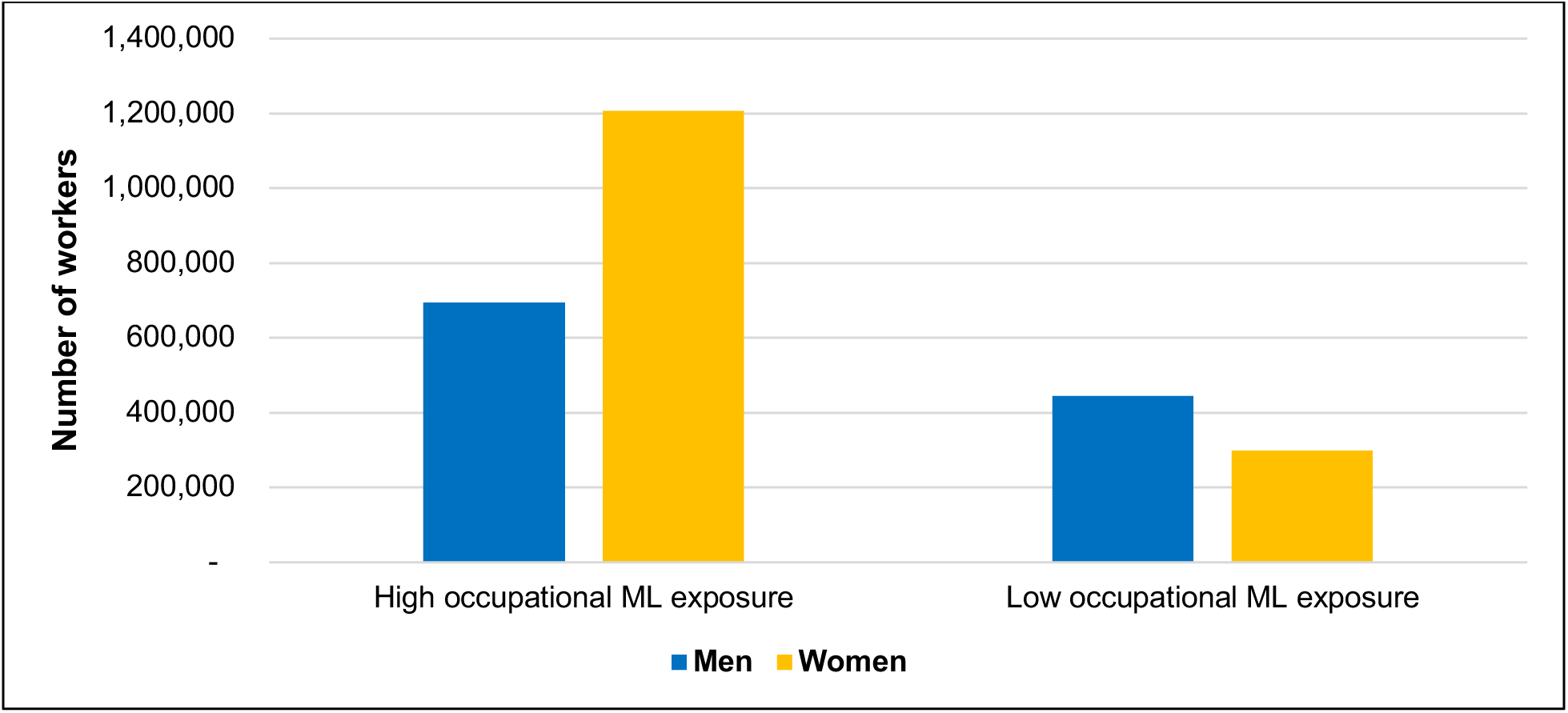
Number Canadian workers with high and low occupational exposure to machine learning (ML)

Table 1 summarizes the descriptive statistics for gender, educational attainment, hourly wages, and job skills, training, and experience requirements according to high or low occupational exposure to ML. Occupations characterized by high ML exposure were more likely to consist of a greater proportion of women workers when compared to men worker (63.4 percent vs. 36.6 percent). In contrast, occupations characterized by low ML exposure were more likely to consist of men compared to women (59.9 percent vs. 40.1 percent).

**Table 1.**
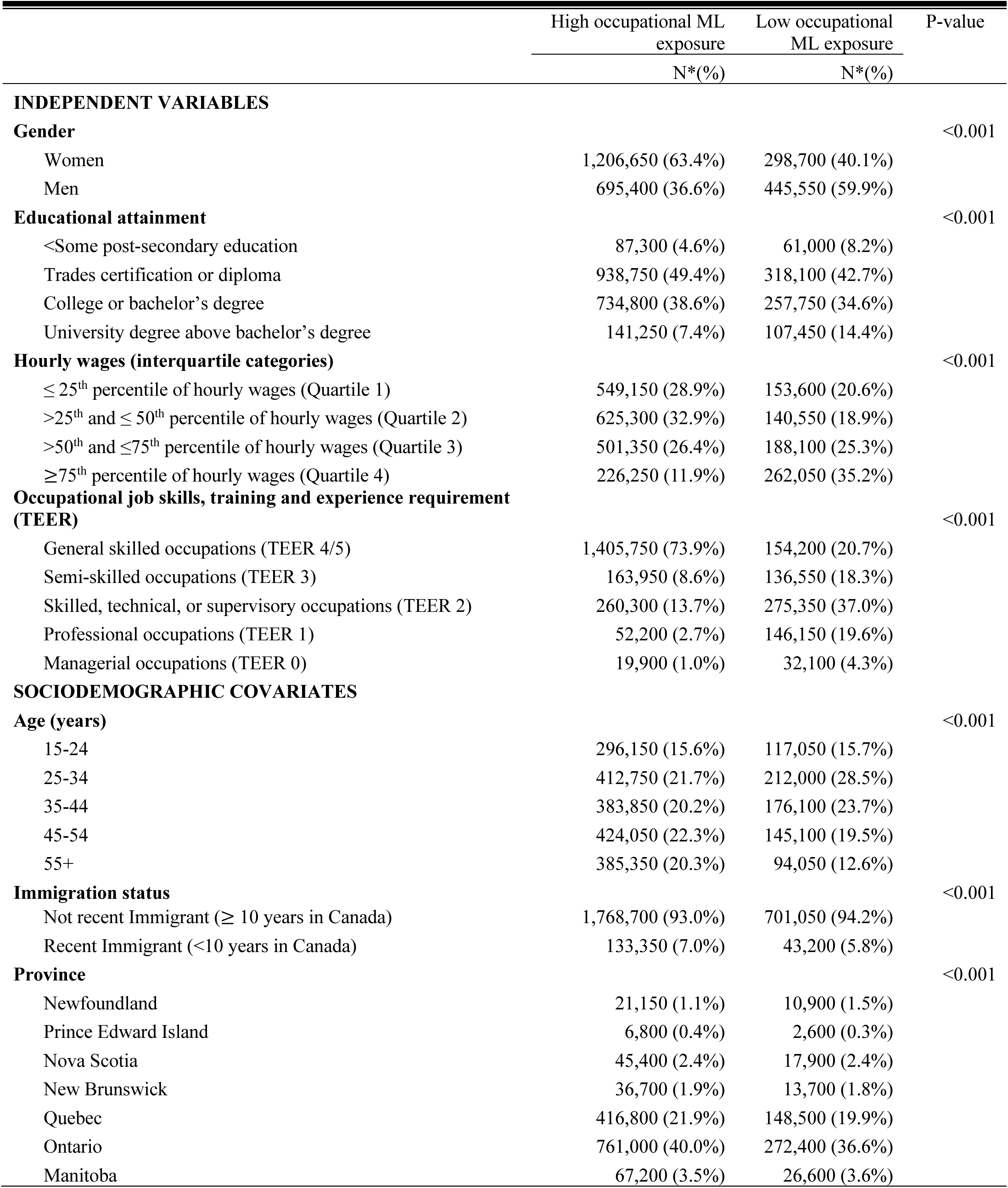
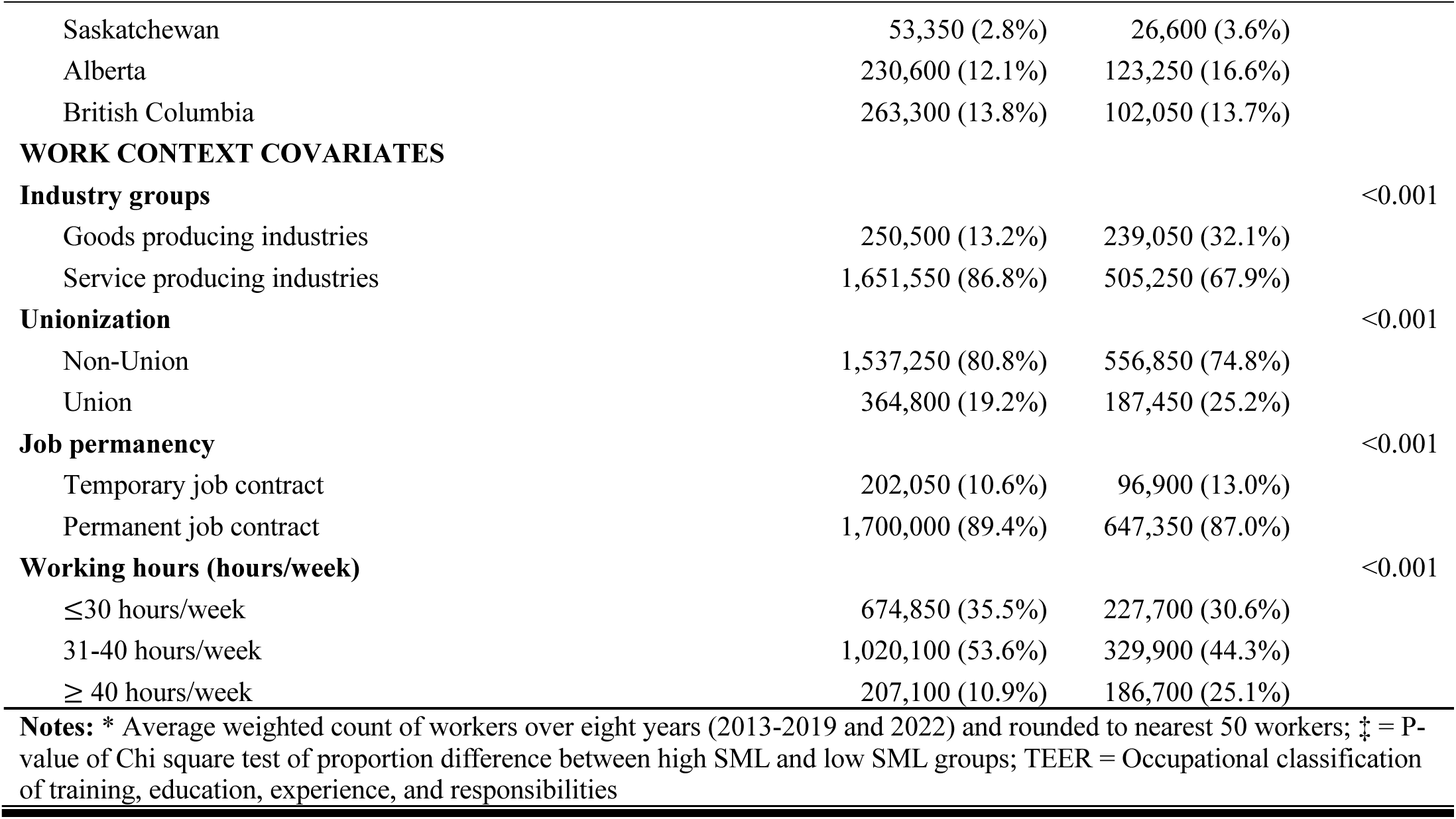
Descriptive statistics of independent variables and study covariates presented based on occupational exposure to machine learning (ML).

|  | High occupational ML exposure | Low occupational ML exposure | P-value |
| --- | --- | --- | --- |
|  | N*(%) | N*(%) |  |
| **INDEPENDENT VARIABLES** |  |  |  |
| **Gender** |  |  | <0.001 |
| Women | 1,206,650 (63.4%) | 298,700 (40.1%) |  |
| Men | 695,400 (36.6%) | 445,550 (59.9%) |  |
| **Educational attainment** |  |  | <0.001 |
| <Some post-secondary education | 87,300 (4.6%) | 61,000 (8.2%) |  |
| Trades certification or diploma | 938,750 (49.4%) | 318,100 (42.7%) |  |
| College or bachelor's degree | 734,800 (38.6%) | 257,750 (34.6%) |  |
| University degree above bachelor's degree | 141,250 (7.4%) | 107,450 (14.4%) |  |
| **Hourly wages (interquartile categories)** |  |  | <0.001 |
| ≤ 25<sup>th</sup> percentile of hourly wages (Quartile 1) | 549,150 (28.9%) | 153,600 (20.6%) |  |
| >25<sup>th</sup> and ≤ 50<sup>th</sup> percentile of hourly wages (Quartile 2) | 625,300 (32.9%) | 140,550 (18.9%) |  |
| >50<sup>th</sup> and ≤ 75<sup>th</sup> percentile of hourly wages (Quartile 3) | 501,350 (26.4%) | 188,100 (25.3%) |  |
| ≥ 75<sup>th</sup> percentile of hourly wages (Quartile 4) | 226,250 (11.9%) | 262,050 (35.2%) |  |
| **Occupational job skills, training and experience requirement (TEER)** |  |  | <0.001 |
| General skilled occupations (TEER 4/5) | 1,405,750 (73.9%) | 154,200 (20.7%) |  |
| Semi-skilled occupations (TEER 3) | 163,950 (8.6%) | 136,550 (18.3%) |  |
| Skilled, technical, or supervisory occupations (TEER 2) | 260,300 (13.7%) | 275,350 (37.0%) |  |
| Professional occupations (TEER 1) | 52,200 (2.7%) | 146,150 (19.6%) |  |
| Managerial occupations (TEER 0) | 19,900 (1.0%) | 32,100 (4.3%) |  |
| **SOCIODEMOGRAPHIC COVARIATES** |  |  |  |
| **Age (years)** |  |  | <0.001 |
| 15-24 | 296,150 (15.6%) | 117,050 (15.7%) |  |
| 25-34 | 412,750 (21.7%) | 212,000 (28.5%) |  |
| 35-44 | 383,850 (20.2%) | 176,100 (23.7%) |  |
| 45-54 | 424,050 (22.3%) | 145,100 (19.5%) |  |
| 55+ | 385,350 (20.3%) | 94,050 (12.6%) |  |
| **Immigration status** |  |  | <0.001 |
| Not recent Immigrant (≥ 10 years in Canada) | 1,768,700 (93.0%) | 701,050 (94.2%) |  |
| Recent Immigrant (<10 years in Canada) | 133,350 (7.0%) | 43,200 (5.8%) |  |
| **Province** |  |  | <0.001 |
| Newfoundland | 21,150 (1.1%) | 10,900 (1.5%) |  |
| Prince Edward Island | 6,800 (0.4%) | 2,600 (0.3%) |  |
| Nova Scotia | 45,400 (2.4%) | 17,900 (2.4%) |  |
| New Brunswick | 36,700 (1.9%) | 13,700 (1.8%) |  |
| Quebec | 416,800 (21.9%) | 148,500 (19.9%) |  |
| Ontario | 761,000 (40.0%) | 272,400 (36.6%) |  |
| Manitoba | 67,200 (3.5%) | 26,600 (3.6%) |  |
| Saskatchewan | 53,350 (2.8%) | 26,600 (3.6%) |  |
| Alberta | 230,600 (12.1%) | 123,250 (16.6%) |  |
| British Columbia | 263,300 (13.8%) | 102,050 (13.7%) |  |
| **WORK CONTEXT COVARIATES** |  |  |  |
| **Industry groups** |  |  | <0.001 |
| Goods producing industries | 250,500 (13.2%) | 239,050 (32.1%) |  |
| Service producing industries | 1,651,550 (86.8%) | 505,250 (67.9%) |  |
| **Unionization** |  |  | <0.001 |
| Non-Union | 1,537,250 (80.8%) | 556,850 (74.8%) |  |
| Union | 364,800 (19.2%) | 187,450 (25.2%) |  |
| **Job permanency** |  |  | <0.001 |
| Temporary job contract | 202,050 (10.6%) | 96,900 (13.0%) |  |
| Permanent job contract | 1,700,000 (89.4%) | 647,350 (87.0%) |  |
| **Working hours (hours/week)** |  |  | <0.001 |
| ≤30 hours/week | 674,850 (35.5%) | 227,700 (30.6%) |  |
| 31-40 hours/week | 1,020,100 (53.6%) | 329,900 (44.3%) |  |
| ≥ 40 hours/week | 207,100 (10.9%) | 186,700 (25.1%) |  |

**Notes:** \* Average weighted count of workers over eight years (2013-2019 and 2022) and rounded to nearest 50 workers; ‡ = P-value of Chi square test of proportion difference between high SML and low SML groups; TEER = Occupational classification of training, education, experience, and responsibilities

The proportion of workers in high and low exposure to ML also differed according to educational attainment. When compared to occupations with low ML exposure, occupations characterized by high exposure to ML were more likely to be composed of workers with a trade’s certification or bachelor’s degree (42.7 percent vs. 49.4 percent) and college or bachelor’s degree (38.6 percent vs. 34.6 percent). When compared to occupations with low ML exposure, occupations characterized by high exposure to ML were less likely to be composed of workers with some post-secondary education or less (8.2 percent vs. 4.6 percent) or a university degree above a bachelor’s degree (14.4 percent vs. 7.4 percent).

When compared to occupations with low exposure to ML, occupations with high exposure to ML were composed with a greater proportion of workers in hourly wage quartile 1 (21 percent vs. 29 percent) and hourly wage quartile 2 (19 percent vs. 33 percent). Workers with low exposure to ML were more likely to consist of workers in hourly wage quartile 4 when compared to occupations with high ML exposure (35 percent vs. 12 percent).

In comparison to occupations characterized by low ML exposure, a greater proportion of high ML exposure occupations were composed of workers in general skilled occupations with the lowest amount of job skills and training requirements (73.9% vs. 20.7%). Occupations characterized by low ML exposure were composed of workers in jobs with greater job skills and training requirements than those with high occupational ML exposure.

Separate gender-stratified multivariable models examined the association between educational attainment (models 1), hourly wages (models 2) and job skills, training, and experiences requirements (models 3) and the odds of high occupational ML exposure when compared to all other Canadian workers (a) or low occupational ML exposure when compared to all other Canadian workers (b) when controlling for study covariates (Table 2). Statistically significant gender differences were identified in all models and key findings will be described in the sections below.

**Table 2.** Multivariable logistic regressions models were fitted to estimate likelihood of employment in high ML exposure occupations and likelihood of employment in Low ML exposure occupation when compared to all other Canadian workers.

| Models | Outcome a: High occupational ML exposure<sup>a</sup> |  |  |  |  |  | Outcome b. Low Occupational ML Exposure<sup>b</sup> |  |  |  |  |  |
| --- | --- | --- | --- | --- | --- | --- | --- | --- | --- | --- | --- | --- |
|  | Women workers |  |  | Men workers |  |  | Women workers |  |  | Men workers |  |  |
|  | OR | Low CI | High CI | OR | Low CI | High CI | OR | Low CI | High CI | OR | Low CI | High CI |
| **Models 1: Educational attainment<sup>‡</sup>** |  |  |  |  |  |  |  |  |  |  |  |  |
| <Some post-secondary education | ref |  |  | ref |  |  | ref |  |  | ref |  |  |
| Trades certification or diploma | 1.091 | 1.074 | 1.110 | 0.517 | 0.507 | 0.527 | 2.272 | 2.208 | 2.338 | 0.993 | 0.975 | 1.010 |
| College or bachelor's degree | 0.917 | 0.909 | 0.925 | 0.862 | 0.853 | 0.871 | 1.268 | 1.244 | 1.291 | 0.924 | 0.911 | 0.937 |
| University degree above bachelor's degree | 0.378 | 0.370 | 0.385 | 0.455 | 0.445 | 0.465 | 1.640 | 1.595 | 1.687 | 0.803 | 0.782 | 0.824 |
| **Models 2. Hourly wages (interquartile categories)<sup>‡</sup>** |  |  |  |  |  |  |  |  |  |  |  |  |
| ≤ 25<sup>th</sup> percentile of hourly wages (Quartile 1) | ref |  |  | ref |  |  | ref |  |  | ref |  |  |
| >25<sup>th</sup> and ≤ 50<sup>th</sup> percentile of hourly wages (Quartile 2) | 1.846 | 1.827 | 1.866 | 0.862 | 0.85 | 0.874 | 0.895 | 0.874 | 0.916 | 1.238 | 1.21 | 1.268 |
| >50<sup>th</sup> and ≤ 75<sup>th</sup> percentile of hourly wages (Quartile 3) | 1.562 | 1.544 | 1.581 | 0.678 | 0.668 | 0.688 | 1.282 | 1.252 | 1.313 | 1.489 | 1.455 | 1.523 |
| >75<sup>th</sup> percentile of hourly wages (Quartile 4) | 0.497 | 0.489 | 0.505 | 0.412 | 0.405 | 0.418 | 1.975 | 1.928 | 2.024 | 2.103 | 2.056 | 2.152 |
| **Model 3. Job skill and training requirements<sup>‡</sup>** |  |  |  |  |  |  |  |  |  |  |  |  |
| General skilled occupation | ref |  |  | ref |  |  | ref |  |  | ref |  |  |
| Semi-skilled occupations | 0.231 | 0.228 | 0.234 | 0.091 | 0.089 | 0.094 | 1.956 | 1.908 | 2.004 | 2.732 | 2.675 | 2.791 |
| Skilled, technical, or supervisory occupations | 0.302 | 0.299 | 0.306 | 0.096 | 0.094 | 0.097 | 1.759 | 1.717 | 1.802 | 2.851 | 2.799 | 2.902 |
| Professional occupations | 0.025 | 0.024 | 0.025 | 0.069 | 0.068 | 0.071 | 2.986 | 2.919 | 3.055 | 1.597 | 1.560 | 1.636 |
| Managerial occupations | 0.026 | 0.025 | 0.028 | 0.062 | 0.060 | 0.064 | 1.765 | 1.700 | 1.832 | 0.856 | 0.825 | 0.889 |
| **Notes:** a = SML score in the top ten percentile compared to the reference group of occupational SML scores outside of the top ten percentile; b = occupations with an SML score in the bottom ten percentile were compared to the reference group of occupational SML scores outside of the bottom ten percentile; OR = odds ratios; CI = confidence interval; Each model adjusted for sociodemographic and work context covariates; <sup>‡</sup> = indicates significant difference between men and women at p <.001. |  |  |  |  |  |  |  |  |  |  |  |  |

For both women and men workers, greater educational attainment was associated with a lower odds of high occupational ML exposure when compared to Canadian working population and when controlling for study covariate (model 1a). Gender-specific effects emerged when examining odds of low occupational ML exposure (model 1b). Women with a university degree above a bachelor’s degree (OR = 1.64, 95%CI, 1.59-1.69), college or bachelor’s degree (OR = 1.27, 95%CI 1.24-1.29) or trades certification or diploma (OR=2.72, 95%CI, 2.21-2.34) had a greater odds of low occupational ML exposure when compared to women with some post-secondary education or less (lowest educational attainment category). Men with a university degree above a bachelor’s degree (OR = 0.80, 95%CI, 0.78-0.82) or a college or bachelor’s degree (OR = 0.92, 95%CI, 0.91-0.94) had a lower likelihood of low ML occupational exposure when compared to men with lowest educational attainment.

**Figure 2.**
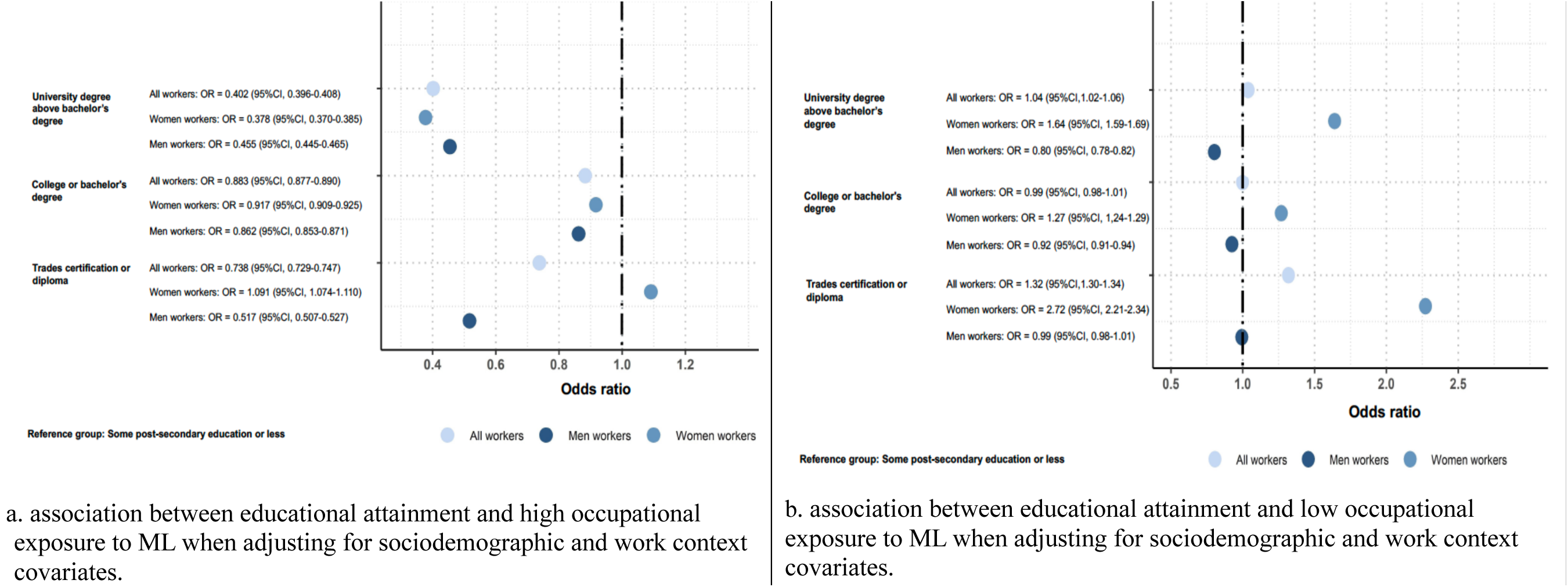
Summary of multivariable logistic regression models examining the association between educational attainment and high occupational machine learning (ML) exposure (a) and low occupational ML exposure (b).

Among both women and men, the highest earners were less likely to make up the workers with high occupational ML exposure (model 2a) and more likely to make up the workers in low ML exposure occupations when controlling for study covariates (model 2b). Of note, women with hourly wages in the 2^nd^ quartile (OR = 1.85, 95%CI 1.83-1.87) and 3^rd^ quartile (OR = 1.56, 95%CI 1.54-1.58) had a greater likelihood of high occupational exposure to ML when compared to lowest wage women earners (quartile 1).

**Figure 3.**
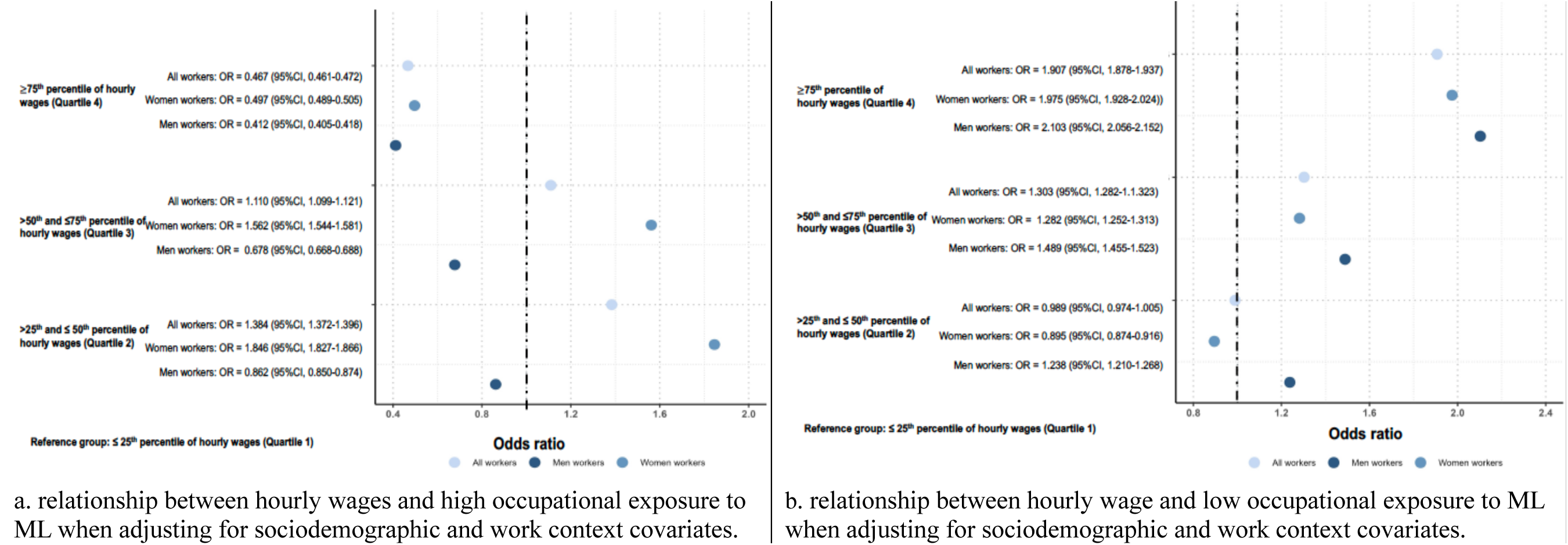
Summary of multivariable logistic regression models examining the relationship between hourly wages and high occupational machine learning (ML) exposure (a) and low occupational ML exposure (b).

Finally, greater job skills, training and experience requirements were associated with a lower likelihood of a high occupational exposure to ML (model 3a) and higher likelihood of reporting low occupational exposure to ML (model 3b). Interestingly, women in managerial positions (with the highest job skills, training, and experience requirements) had a greater odds of low ML exposure when compared to women workers in general skilled occupations (OR = 1.77, 95CI% 1.70-1.83). Men workers in managerial occupations were less likely to be low ML exposure when compared to those with general skilled occupations (OR = 0.86, 95%CI 0.83-0.89).

**Figure 4.**
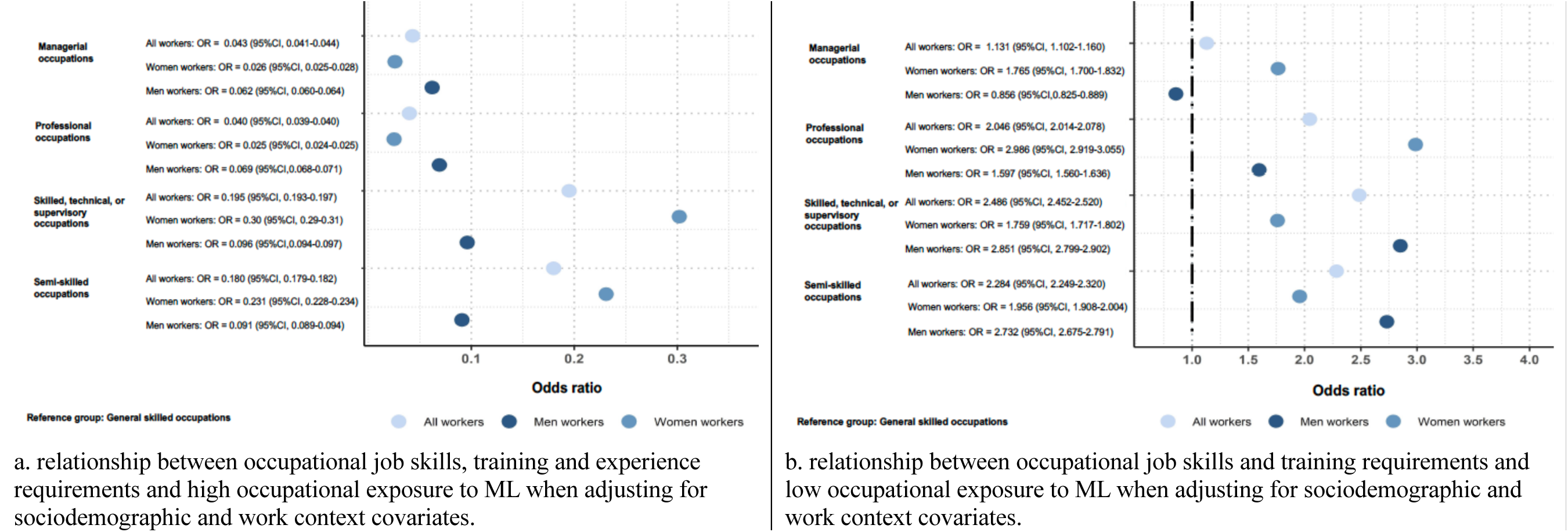
Summary of multivariable logistic regression models examining the relationship between occupational job skills and training requirements and high occupational machine learning (ML) exposure (a) and low occupational ML exposure (b).

## DISCUSSION

The adoption of ML within Canadian workplaces has the potential to drive rapid change to the nature and availability of work and may also exacerbate workforce inequities that can disrupt access to critical social determinants of health. Our study is among the first we are aware of that provides a snapshot of occupational ML exposure in Canada’s labour market and the extent to which high and low occupational ML exposure may be associated with worker sociodemographic and occupational characteristics. Like other technological transformations that have shaped Canada’s labour market, we show that historically disadvantaged segments of the workforce may be most likely to have their occupations affected by ML. We also show that ML may have a gendered effect and disproportionately impact women when compared to men. Findings can inform targeted policies and programs that optimize the benefits of ML and address the potential adverse effects of the technology on workers, especially amongst groups with the highest occupational exposure.

We utilized a novel analytical approach to describe Canadian occupations based on the extent to which they are made up of job tasks that are suitable to being performed by ML. Through this approach we estimated the number of workers in occupations with the highest (i.e., occupations most suitable to being performed by ML) and lowest exposure to ML (i.e., occupations least suitable to being performed by ML). Through our analytical approach we show that close to two million Canadian workers are employed in occupations with high exposure to ML, which represents close to 11.5 percent of the overall workforce. Less than half of that number of Canadian workers were in occupations characterized by low ML exposure (744,250 workers representing 4.5 percent of the Canadian workforce). It is not clear based on our analytical approach whether workers in jobs with high exposure to ML will experience advantage or disadvantage stemming from the technology. Drawing from past research there are several ways that our findings could be interpreted. First, workers in occupations with high exposure to ML could be at risk of partial or full job displacement. Second, the use of ML may complement the work performed by workers in high exposure jobs and free up time for high value activities and productivity gains ^4^ ^32^ ^33^. On the other hand, while those with low occupational ML exposure are less likely to work in a job where they face less of a risk of being displaced by a machine, they may also be less likely to leverage benefits associated with ML to the performance of their job tasks. Additional research is needed to expand on our results and specifically examine how ML both positively and negatively impacts the employment and working conditions for different groups of workers.

We also found that all Canadian occupations consisted of job tasks that could be performed by ML. Findings align with past studies in Canada and the US which indicate that the technology can have a widespread impact on the economy ^4^ ^22^. At the same time, our study show that no single occupation could be completely suitable to being performed by ML and the ability for machines to match humans in all job tasks remains limited ^5^. Findings support an ongoing perspective that ML will contribute to changes to Canadian workplaces and the role of workers. There is a need to prepare all workers for the growing adoption of ML and other AI so that they can work alongside technology. The importance of studying the impact of ML on the nature of work will continue to grow as the technology advances and gains greater independence and surpasses human learning and reasoning ^34–36^.

Scholars have previously posited that the adoption of ML within the labour will have a disproportionate impact on workers in higher-skilled and better paying occupations whose jobs tasks require a greater degree of prediction ^9^ ^17^ ^37^. Our results, however, align with research of past waves of automation and show high occupational ML exposure among workers with less educational attainment or those in lower-paying occupations and occupations with fewer job training, skill, and experience requirements. Our study signals a potential risk that the adoption of ML could pose to the labour market by reinforcing or even widening inequities. Of particular concern, ML has the potential to contribute to wage polarization between groups of workers with high and low exposure to ML. The transfer of work responsibilities from workers to machines is expected to lower the cost of labour and contribute to a downward pressure on wages which could disadvantage vulnerable worker groups ^4^ ^37^. Alternatively, ML may be used to perform the most repetitive or unsafe job tasks and contribute to working conditions that are more optimal to groups of ^38^ ^39^. Additional research is needed to elaborate on our study findings and build a better understanding of the mechanisms by which ML may positively or negatively impact working conditions and different groups of workers to develop targeted policy interventions and supports.

Occupational exposure to ML in the Canadian labour market may also be patterned according to gender. Men and women tend to cluster in different occupations which shape their employment experiences and exposure to different technologies in the workplace ^13^ ^40^. Our findings add to the evidence on gender segregation in the labour market related to technological transformations. We show that Canadian women may be employed in occupations where their tasks are most suitable to ML and may be more likely to experience high exposure to ML ^3^. The implications of these findings could be interpreted in several ways. The job tasks that women workers perform and the job roles they hold may be at a greater risk of being substituted by ML. Alternatively, women may also be more likely to benefit from ML including the technology’s ability to complement their tasks and contribute to productivity gains. At the same time, our findings may suggest that men may be shielded from job displacement related to ML. Men may also be more likely to be excluded from the economic opportunities that can emerge because of ML adoption. Findings highlight the importance of future research to unpack how ML adoption may impact men and women differently and the potential need for gender-sensitive policy and programmatic approaches to address the challenges and opportunities of ML for Canadian workers.

To further unpack gender differences, our study examined whether the relationship between educational attainment, hourly wages and job skills, training and experience requirements and ML exposure differed for women and men. Interestingly, our findings indicated that greater educational attainment and being in a managerial occupation was associated with low occupational ML exposure among women. Past studies have indicate that educational attainment may be particularly important among women and insulate them from economic challenges including automation risk ^9^ ^41^. At the same time, our study could suggest that highly educated women and those in management positions may be excluded from ML-related economic opportunities that could reinforce or widen gender divides. Future research is required to better understand the how ML may impact the employment experiences of men and women in the labour market and how differences may stem from worker and occupational characteristics.

Our study has several strengths and limitations. We use a novel analytical approach that measures the extent to which different Canadian occupations are made up of job tasks that are suitable to being performed by ML. We use eight cycles of Canada’s LFS to produce a population-based estimates of ML exposure. We were also able to describe occupations with high and low ML exposure according to occupational and worker characteristics. Findings provide an important description of the extent to which ML may impact Canada’s labour market and the ways in which the technology could contribute to labour market segregation. There are also some limitations to note. Our measure focuses on the technical feasibility of ML and its suitability to perform job tasks ^4^ ^22^. We are unable to comment on how ML is used in practice and the specific ways in which it will impact workers and the way they perform their jobs. Our findings do not elaborate on the economic, organizational, legal, cultural, and societal factors that may facilitate or inhibit ML adoption within the labour market and its impact on occupations and workers.

While we utilize an established measure of occupational ML exposure, other measures exist including those that capture AI exposure at the industry or geographic levels ^42^ as well those which examine the extent to which AI may be used to complement or substitute workers ^32^. Moreover, the measure we selected does not evaluate all forms of AI and the automation of work such as those driven by the most recent advances in generative AI ^25^. Recent measures have been developed to examine the occupational impact of generative AI ^25^. These measures are preliminary and require further validation before being more widely used for labour market estimates. Consequently, ongoing research is required to examine how emerging forms of AI will impact workers and workplaces.

## CONCLUSION

ML, an AI subfield, has the potential to transform Canada’s labour market and contribute to labour market and health inequities. Our analysis of labour force data offers a critical picture of the occupations most affected by ML and indicate that a substantial number of Canadians are working in jobs where they may be likely to be exposed to ML. Workers in jobs with high exposure to ML may face both challenges and opportunities associated with the technology. We also highlight that occupational ML exposure may contribute to the segregation of the labour market according to educational attainment, wages and occupational job skills and training requirements. Our study provides a foundational evidence base that can be used to direct attention and efforts towards occupations and worker groups who may be most likely to directly be exposed to the ML to develop strategic responses that ensure inclusion in a working world where ML and other forms of AI is being increasingly used.

## Data Availability

All data produced in the present work are contained in the manuscript

## Acknowledgements

The study team would like to acknowledge the contributions of Marc Frenette from Statistics Canada and Michael Willcox from the Labour Market Information Council who provided analytical expertise to the project. We would also like to thank Hela Bakhtari for project coordination support.

## FUNDING STATEMENT

The study was supported by Social Sciences and Humanities Research Council of Canada Partnership Development Grant (#890-2021-0018), and the Future Skills Centre (#20220524). Dr. Arif Jetha’s salary is partially supported by a Stars Career Development Award from the Arthritis Society (Canada). Funding bodies had no role in study design or manuscript writing.

## COMPETING INTERESTS

AJ, QL, FVS, AB and PS are employees of the Institute for Work & Health (IWH), which is supported through funding from the Ontario Ministry of Labour, Immigration, Training and Skills Development (MLITSD). The analyses, conclusions, opinions, and statements expressed herein are solely those of the authors and do not reflect those of the MLITSD; no endorsement is intended or should be inferred.

**Appendix 1.**
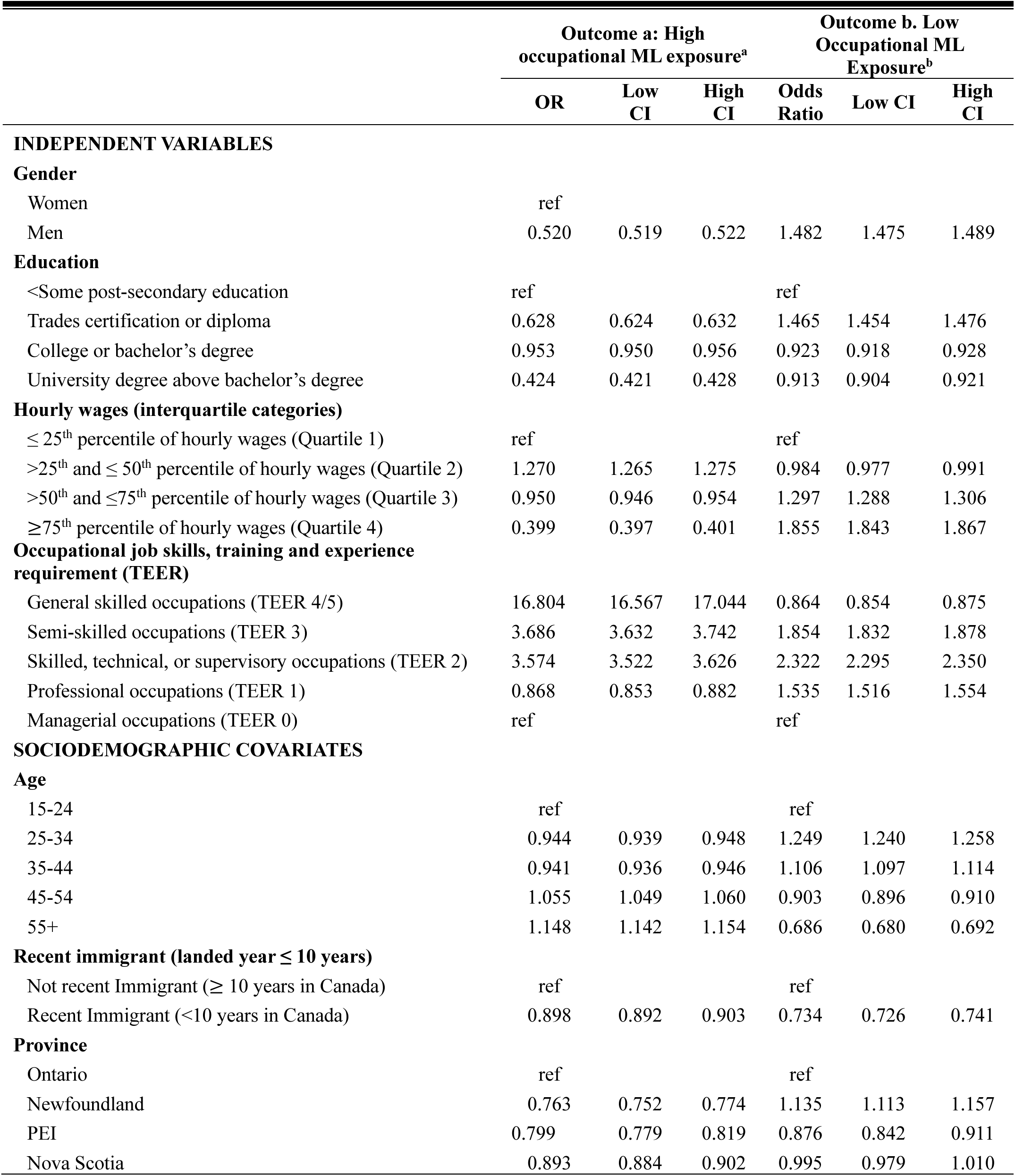
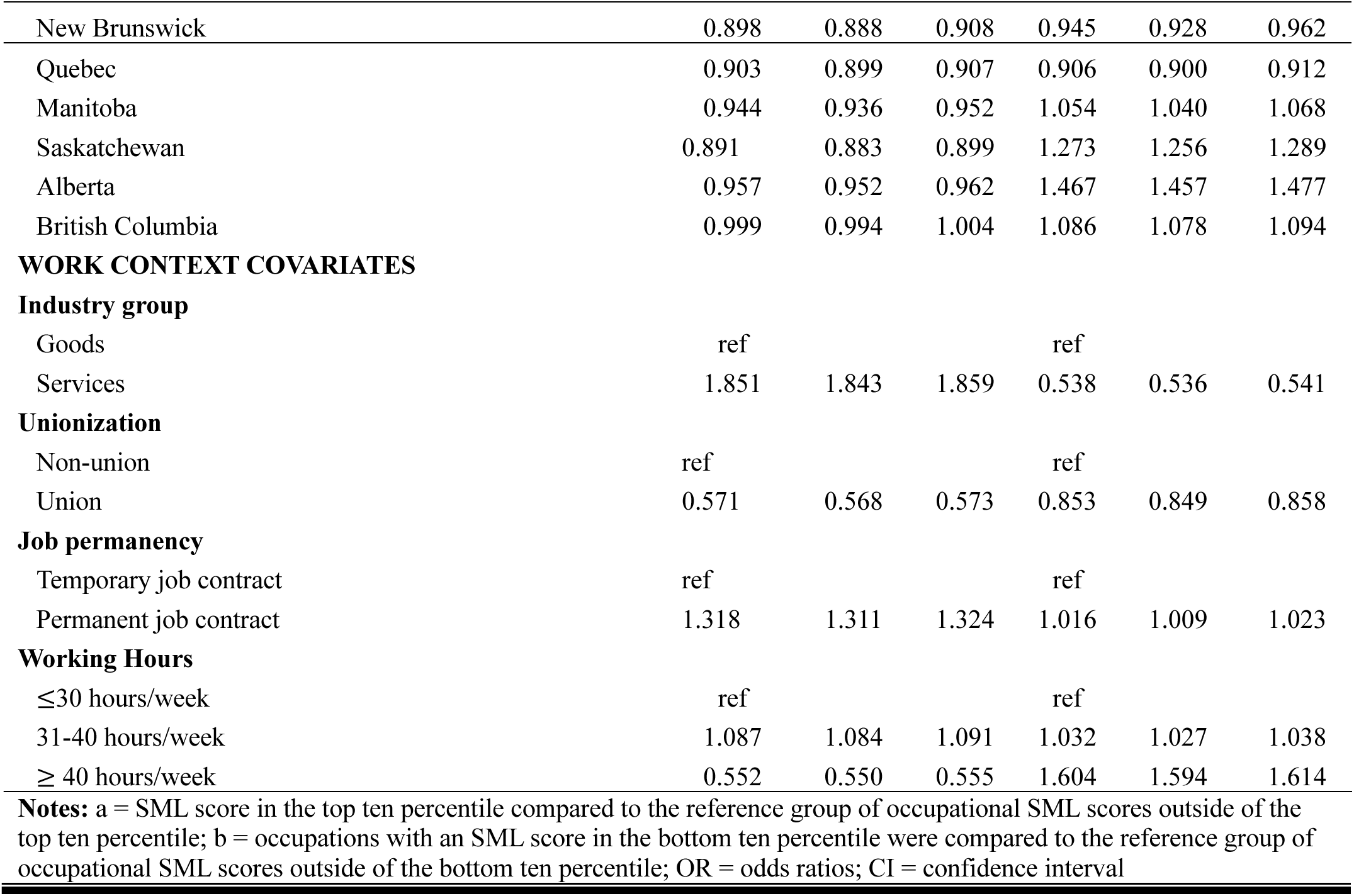
Univariate models estimating likelihood of employment in high ML exposure occupations and likelihood of employment in low ML exposure occupation when compared to all other Canadian workers.

**Appendix 2.**
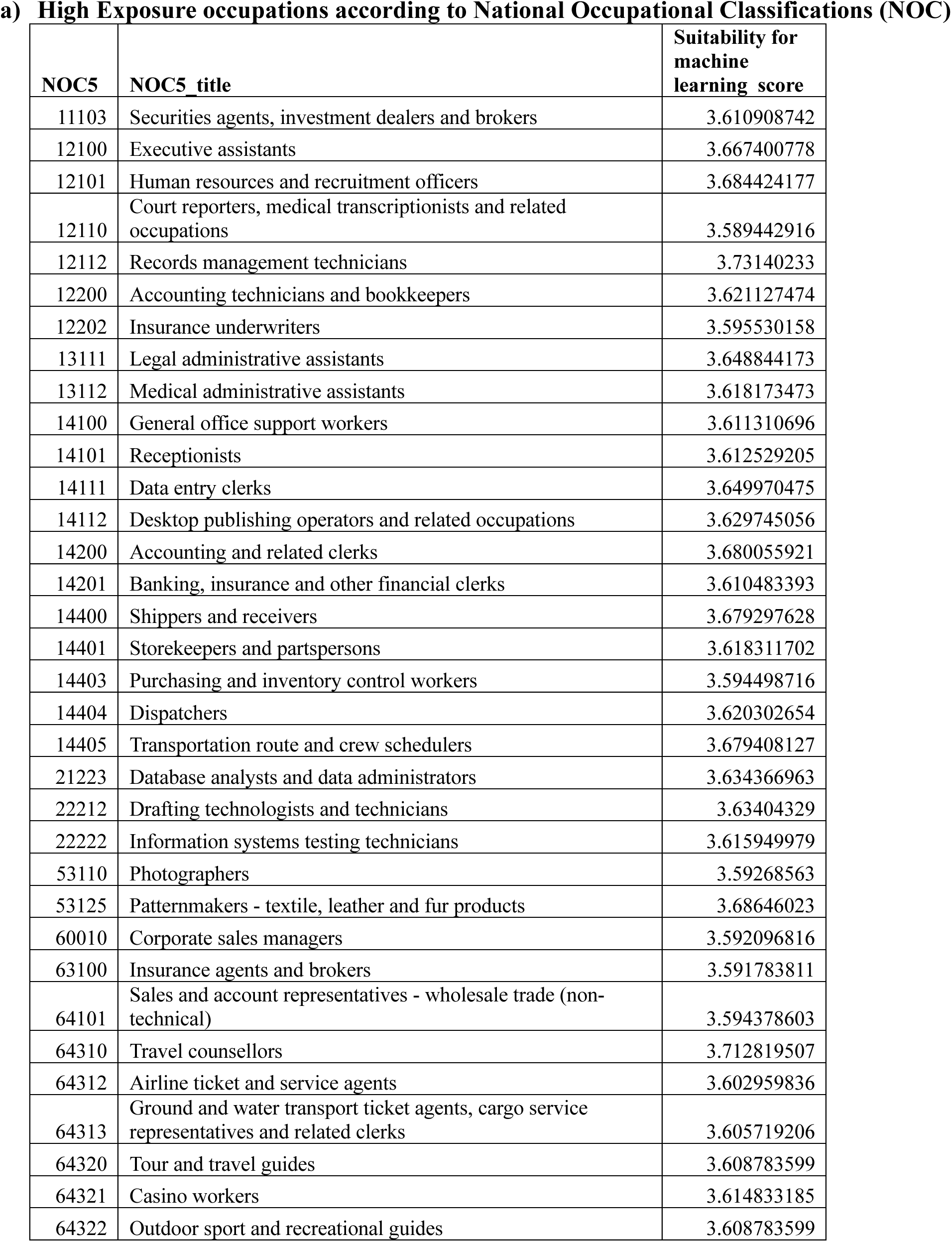
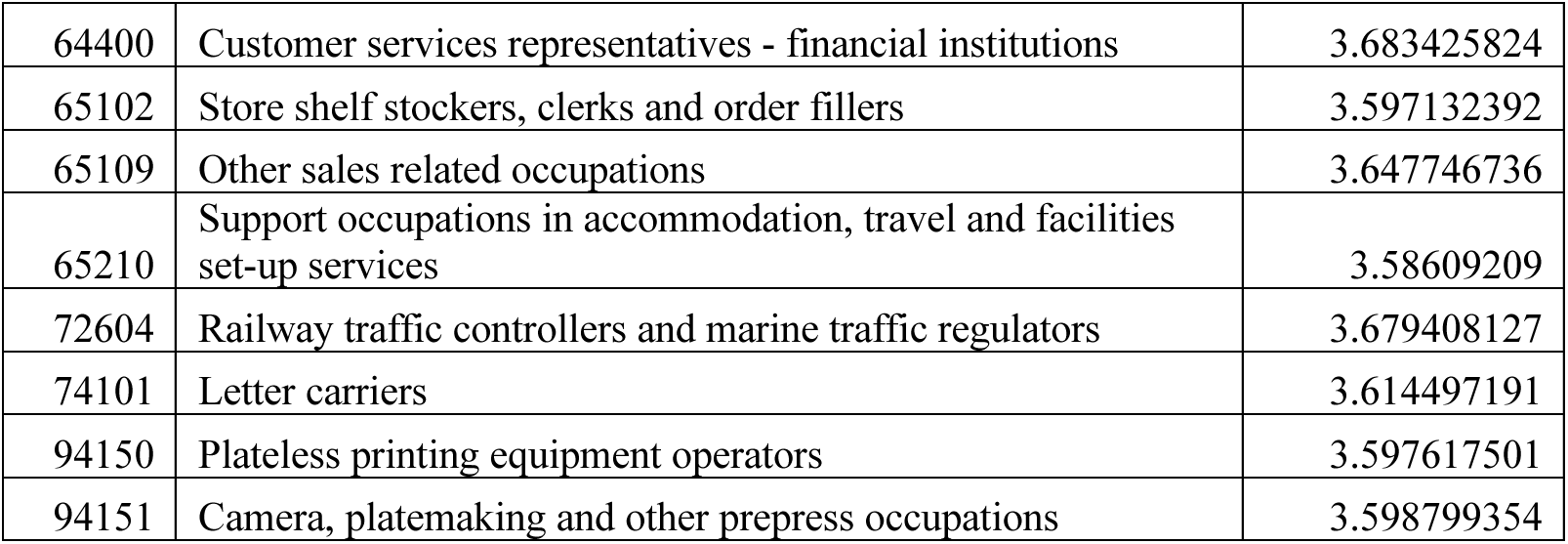
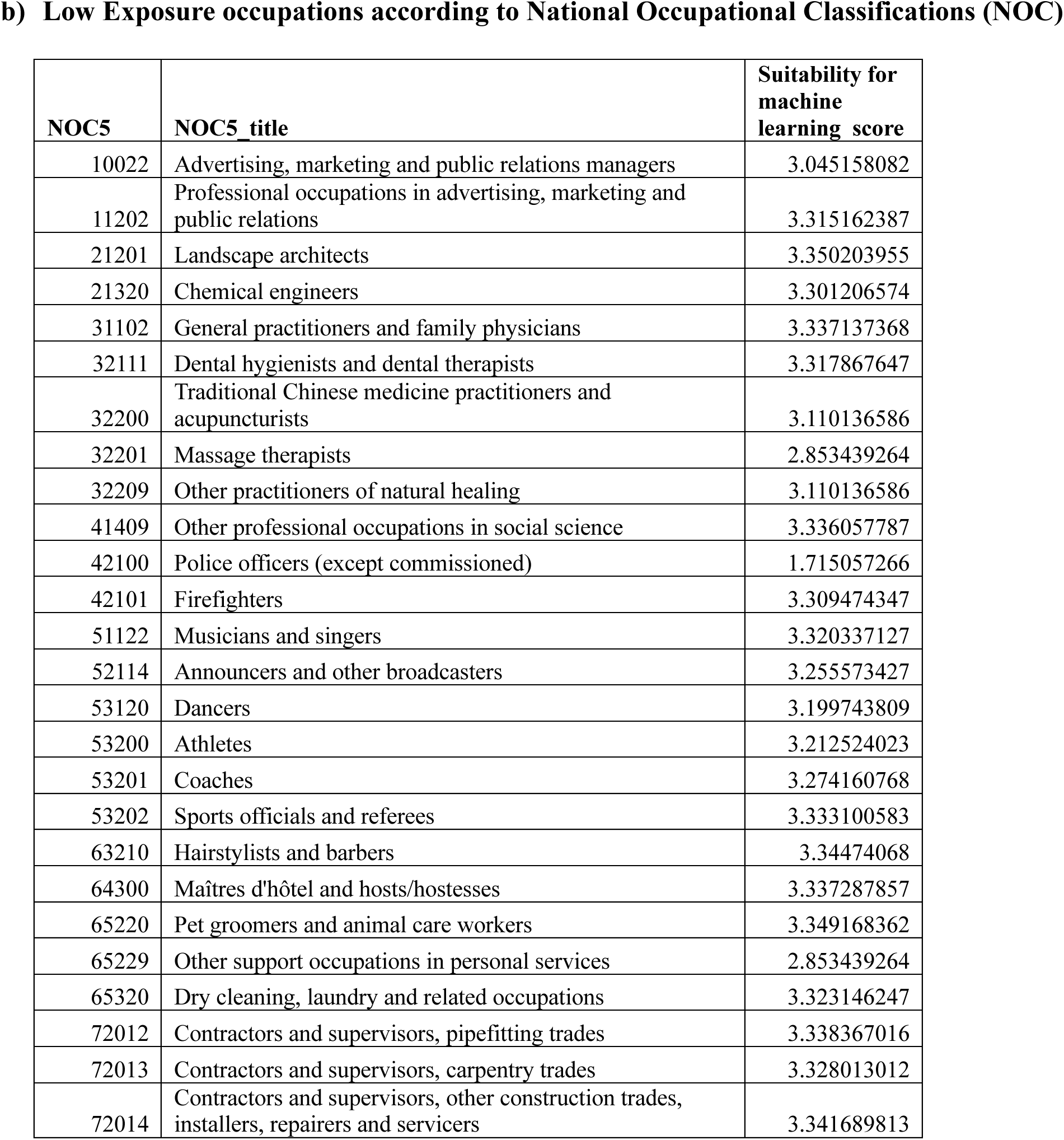
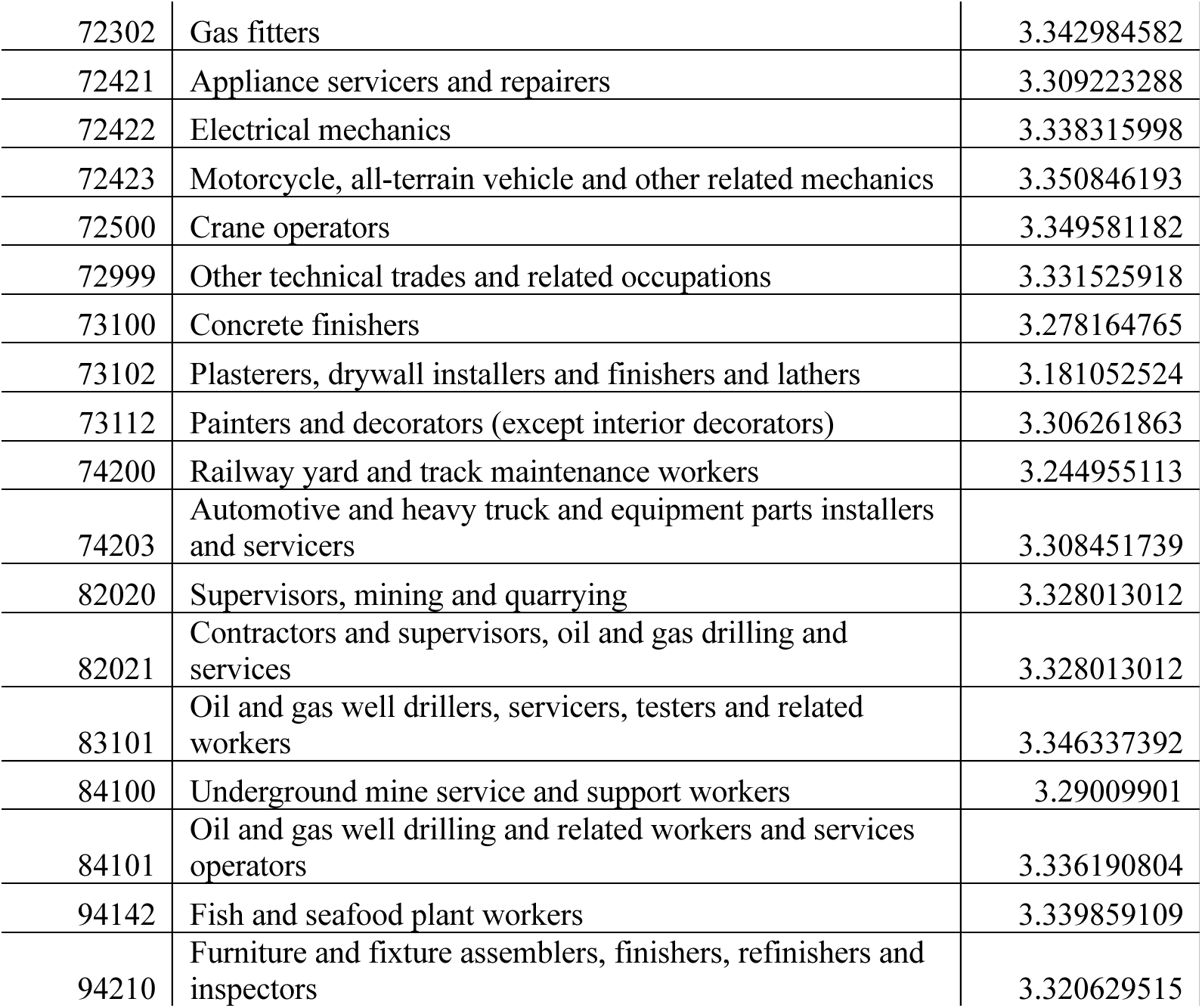
Canadian occupations with high and low machine learning (ML) exposure.

**Appendix 3.**
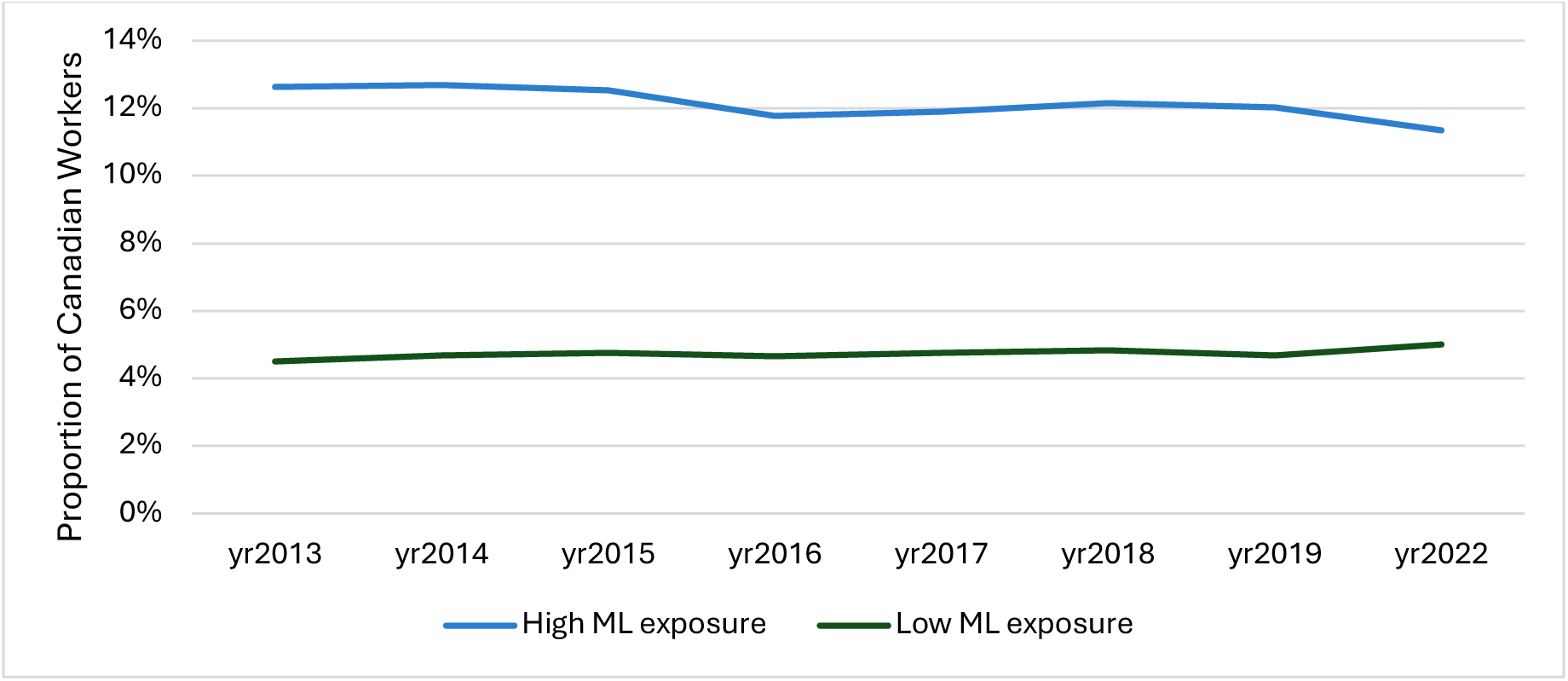
Proportion of Canadian workers in occupations with high and low exposure to machine learning (ML) over the eight-year study period.

## Notes

### Competing Interest Statement

The authors have declared no competing interest.

### Author Declarations

No ethics review was required for this study using secondary Labour Force Survey data. The Labour Force Survey data for this study can only be accessed with security clearance and access to a Statistics Canada Research Data Centre.

## REFERENCES

1. Frank J, Mustard C, Smith P, et al. Work as a social determinant of health in high-income countries: past, present, and future. The Lancet 2023;402(10410):1357–67.

2. Block S, Galabuzi-Grace E, Tranjan R. Canada’s colour coded income inequality: Canadian Centre for Policy Alternatives Ottawa 2019.

3. Quinn EK, Harper A, Rydz E, et al. Men and women at work in Canada, 1991–2016. Labour & Industry: a journal of the social and economic relations of work 2020;30(4):401–12.

4. Brynjolfsson E, Mitchell T. What can machine learning do? Workforce implications. Science 2017;358(6370):1530–34. doi: 10.1126/science.aap8062

5. Brynjolfsson E, Mitchell T, Rock D. What can machines learn, and what does it mean for occupations and the economy? AEA Papers and Proceedings 2018;108:43–47. doi: 10.1257/pandp.20181019

6. Autor D, Dorn D. The growth of low-skill service jobs and the polarization of the US labor market. Am Econ Rev 2013;103(5):1553–97. doi: 10.1257/aer.103.5.1553

7. Jetha A, Shamaee A, Bonaccio S, et al. Fragmentation in the future of work: a horizon scan examining the impact of the changing nature of work on workers experiencing vulnerability. Am J Ind Med 2021;64(8):649–66. doi: 10.1002/ajim.23262 [published Online First: 2021/06/14]

8. Lamb C. The talented Mr. Robot: the impact of automation on Canada’s workforce Toronto (ON): Brookfield Institute for Innovation+ Entrepreneurship; 2016 [Available from: https://brookfieldinstitute.ca/the-talented-mr-robot/ accessed January 22 2023.

9. Acemoglu D, Restrepo P. Low-skill and high-skill automation. J Hum Cap 2018;12(2):204–32. doi: 10.1086/697242

10. OECD. Education at a glance 2022: OECD indicators: OECD Paris 2022.

11. Frey CB, Osborne M. The future of employment. 2013

12. Devillard S, Bonin G, Madgavkar A, et al. The present and future of women at work in Canada. Women Matter: McKinsey, 2019:1–92.

13. Madgavkar A, Manyika J, Krishnan M, et al. The future of women at work: Transitions in the age of automation McKinsey Global Institute, 2019:1–168.

14. Baboolall D, Pinder D, Stewart III S. How automation could affect employment for women in the United Kingdom and minorities in the United States. The McKinsey Quarterly 2019

15. Cortes GM, Jaimovich N, Siu HE. The "end of men" and rise of women in the high-skilled labor market: National Bureau of Economic Research, 2018.

16. Dabla-Norris E, Kochhar K. Women, technology, and the future of work 2018 [Available from: https://www.imf.org/en/Blogs/Articles/2018/11/16/blog-Women-Technology-the-Future-of-Work#:~:text=Digitalization%2C%20artificial%20intelligence%2C%20and%20machine,to%20break%20the%20glass%20ceiling.

17. Webb M. The impact of artificial intelligence on the labor market. SSRN 2019:61. doi: 10.2139/ssrn.3482150

18. Brown S. Machine learning, explained 2021 [Available from: https://mitsloan.mit.edu/ideas-made-to-matter/machine-learning-explained accessed November 1 2023.

19. Li W, Jia F, Hu Q. Automatic segmentation of liver tumor in CT images with deep convolutional neural networks. Journal of Computer and Communications 2015;3(11):146–51.

20. Yee OS, Sagadevan S, Malim NHAH. Credit card fraud detection using machine learning as data mining technique. Journal of Telecommunication, Electronic and Computer Engineering (JTEC) 2018;10(1-4):23–27.

21. Navarro PJ, Fernandez C, Borraz R, et al. A machine learning approach to pedestrian detection for autonomous vehicles using high-definition 3D range data. Sensors 2016;17(1):18.

22. Amarasinghe U, Han F, Oschinski M, et al. Automation affecting Canadians: Understanding the impact of machine learning on the Canadian labour force. Toronto, ON: MaRS Discovery District, 2020:1–25.

23. Kochhar R. Which US Workers Are More Exposed to AI on Their Jobs? 2023

24. Future of jobs report 2023. World Economic Forum, Geneva, Switzerland https://wwwweforumorg/reports/the-future-of-jobs-report-2023; 2023.

25. Eloundou T, Manning S, Mishkin P, et al. Gpts are gpts: An early look at the labor market impact potential of large language models. arXiv preprint arXiv:230310130 2023

26. Statistics Canada. Labour Force Survey (LFS) 2024 [Available from: https://www23.statcan.gc.ca/imdb/p2SV.pl?Function=getSurvey&SDDS=3701 accessed March 1 2024.

27. Statistics Canada. COVID-19 in Canada: A one-year update on social and economic impacts, 2021.

28. U.S. Department of Labor. O*NET 2021 [Available from: https://www.dol.gov/agencies/eta/onet accessed November 1 2023.

29. U.S. Bureau of Labor Statistics. Standard Occupational Classification 2018 [Available from: https://www.bls.gov/soc/2018/home.htm accessed November 1 2023.

30. Statistics Canada. National Occupational Classification (NOC) 2021 Version 1.0 2021 [Available from: https://www23.statcan.gc.ca/imdb/p3VD.pl?Function=getVD&TVD=1322554&adm=0&dis=0 accessed Novmber 1 2023.

31. Allison P. Logistic regression using SAS®: Theory and application. Second Edition ed. Cary, NC, 2012.

32. Pizzinelli C, Panton AJ, Tavares MMM, et al. Labor market exposure to AI: Cross-country differences and distributional implications: International Monetary Fund 2023.

33. Brynjolfsson E, Rock D, Tambe P. How Will Machine Learning Transform the Labor Market? Governance in an Emerging New World 2019;619

34. Goldman Sachs. Generative AI could raise global GDP by 7% New York (NY): Goldman Sachs; 2023 [Available from: https://www.goldmansachs.com/intelligence/pages/generative-ai-could-raise-global-gdp-by-7-percent.html accessed June 6 2023.

35. Agrawal A, Gans J, Goldfarb A. Prediction machines: the simple economics of artificial intelligence. Harvard Way, BA: Harvard Business Press 2018.

36. Russell S. Human-Compatible Artificial Intelligence, 2022.

37. Acemoglu D, Restrepo P. Tasks, automation, and the rise in us wage inequality. Econometrica 2022;90(5):1973–2016.

38. Manyika J, Lund S, Chui M, et al. Jobs lost, jobs gained: workforce transitions in a time of automation, 2017:1–23.

39. Acemoglu D, Restrepo P. Artificial intelligence, automation and work (NBER working paper series no. 24196). National Bureau of Economic Research 2018 doi: 10.3386/w24196

40. Collett C, Gomes LG, Neff G. The effects of AI on the working lives of women: UNESCO Publishing 2022.

41. Moyser M. Women and Paid Work. Women in Canada: A gender-based statistical report. Ottawa, ON: Statistics Canada, 2017:1–38.

42. Felten E, Raj M, Seamans R. Occupational, industry, and geographic exposure to artificial intelligence: A novel dataset and its potential uses. Strategic Management Journal 2021;42(12):2195–217.

